# Factors Influencing Vitamin D Status in Guiyang, China: A Random Forest and SHAP Analysis

**DOI:** 10.64898/2026.05.13.26353105

**Authors:** Pan Ben-you, Wu Xian-ding, Yu Hong-lan

## Abstract

**Objective:** To assess serum 25-hydroxyvitamin D [25(OH)D] levels in a health examination population in Guiyang, a low-latitude, high-altitude, and cloudy city in southwestern China, and to identify key determinants using machine learning.

**Methods:** This retrospective study included 10,931 adults (>20 years) who underwent health checkups at Guiyang First People’s Hospital between February 2019 and April 2025. Beyond conventional statistical comparisons, a two-stage machine learning approach was applied: LASSO regression for feature selection, followed by an optimized Random Forest regression model (mtry = 2). SHapley Additive exPlanations (SHAP) were used to quantify variable importance.

**Results:** The median serum 25(OH)D level was 36.63 (IQR 24.77,53.17) nmol/L. Vitamin D deficiency (<50 nmol/L) was present in 70.98% of participants, while sufficiency (>75 nmol/L) was only 7.35%. Significantly lower levels were observed in females, in adults aged <30 years (deficiency rate 85.6%), and during spring. The optimized Random Forest model achieved a cross-validated RMSE of 21.427. SHAP analysis revealed a clear hierarchy of importance: age (mean SHAP = 5.604) > season (4.104) > sex (1.533) ≈ BMI (1.501).

**Conclusion:** Vitamin D deficiency is highly prevalent in the Guiyang health examination population. Age and season are the dominant determinants, far outweighing sex and BMI. Targeted interventions should focus on young adults, females, and the spring season, especially in regions with similar cloudy highland climates.

## Introduction

Vitamin D is an essential fat-soluble vitamin. Its major circulating form, 25(OH)D, not only plays a central role in calcium-phosphate metabolism and bone health but has also been linked to the risk of various chronic diseases [1–5]. Human vitamin D status depends primarily on cutaneous synthesis under ultraviolet (UV) exposure, making it highly susceptible to geographic climate and lifestyle factors.

Guiyang, a typical low-latitude (26°N) plateau mountain area with an average altitude of approximately 1100 m, is well known for its cloudy and rainy weather—”no three consecutive sunny days.” Annual sunshine hours are short, and actual UV exposure may be severely limited. This unique climatic and geographic setting may place local residents at a substantially higher risk of vitamin D deficiency than other regions. Globally, vitamin D deficiency has been recognized as a pandemic, affecting an estimated 30–60% of children and adults worldwide, with inadequate sun exposure being the major cause . Very few foods naturally contain vitamin D, making cutaneous synthesis under UVB radiation the primary source for most populations [6].However, large-scale epidemiological data on vitamin D status in the general population of Guiyang remain scarce, hampering the development of locally tailored public health interventions.

To address this gap, we retrospectively analyzed serum 25(OH)D levels in 10,931 individuals undergoing health checkups at Guiyang First People’s Hospital. By combining conventional statistics with interpretable machine learning, we aimed to characterize the current vitamin D status and to identify its key determinants, thereby providing a scientific basis for targeted nutrition interventions.

## Methods

### Study population

A total of 10,931 adults (4,273 men, 6,658 women) aged 20–96 years who underwent health examinations at Guiyang First People’s Hospital from February 2019 to April 2025 were included. Participants were categorized into six age groups: <30, 30–40, 40–50, 50– 60, 60–70, and >70 years. Seasons were defined as spring (March–May), summer (June– August), autumn (September–November), and winter (December–February) based on local meteorological conventions.

### Laboratory measurement

Serum 25(OH)D levels were measured using a Siemens ADVIA Centaur XP fully automated chemiluminescence immunoassay system with dedicated reagents. Daily internal quality control and annual external quality assessment (organized by the National Center for Clinical Laboratories, China) were performed to ensure accuracy and reliability.

### Definitions of vitamin D status

According to the Chinese consensus on clinical application of vitamin D and its analogs [7], vitamin D status was categorized as follows: sufficiency >75 nmol/L, insufficiency 50– 75 nmol/L, and deficiency <50 nmol/L.

### Statistical analysis

Conventional statistical analyses were performed using SPSS 31.0. Non-normally distributed continuous data were expressed as median (interquartile range). Comparisons between two groups used the Mann-Whitney U test, and among multiple groups the Kruskal-Wallis H test. Categorical data were compared using the chi-square test. A P value <0.05 was considered statistically significant.

### Machine learning modeling

The analysis was conducted using EasyR online software. The total sample was randomly split into a training set (n = 8,744) and a testing set (n = 2,187). An optimized Random Forest regression model was built with the parameter setting mtry = 2 (number of variables considered at each split). Model performance was assessed using root-mean-square error (RMSE), coefficient of determination (R^2^), and mean absolute error (MAE). Feature importance was interpreted via SHAP values.

## Results

### Baseline characteristics

Detailed baseline characteristics of the study population, including height, weight, blood counts, liver and kidney function, lipids, and glucose, are provided in Supplementary Table S1.

### Distribution of serum 25(OH)D levels

The serum 25(OH)D levels of the entire cohort were 36.63 (24.77, 53.17) nmol/L, with a log-normal distribution (skewness 1.776, kurtosis 7.081). According to the clinical consensus, 7.35% (803/10,931) were vitamin D sufficient, 21.67% (2,369/10,931) insufficient, and 70.98% (7,759/10,931) deficient.

### Vitamin D status by sex, age, and season

Table 1 presents the median 25(OH)D levels and deficiency rates stratified by sex, age group, and season.

**Table 1.**
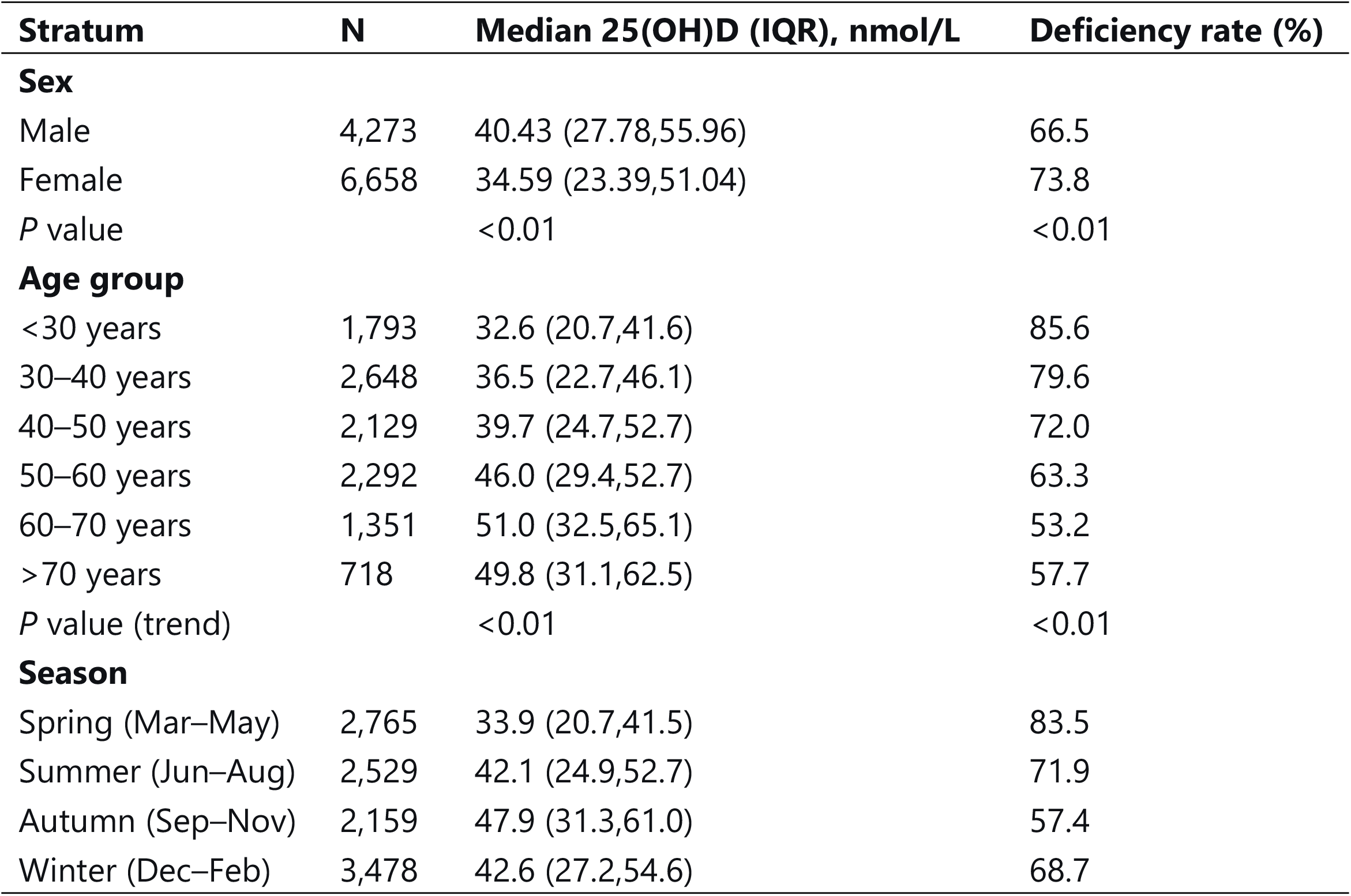
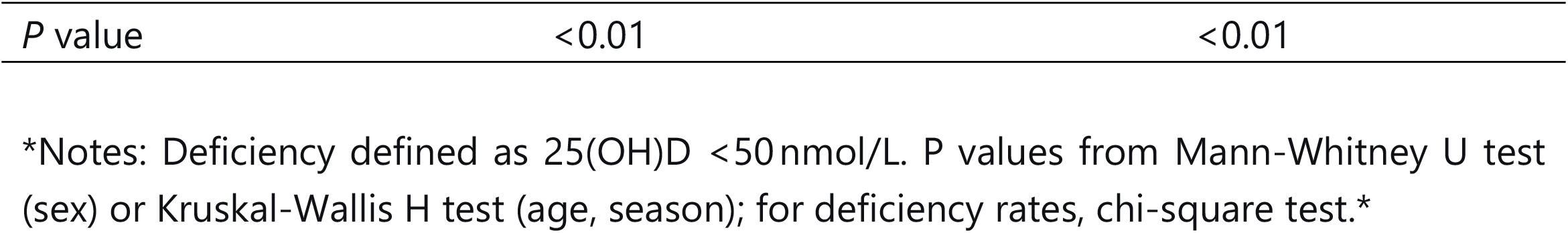
Serum 25(OH)D levels and vitamin D deficiency rates by sex, age group, and season

| Stratum | N | Median 25(OH)D (IQR), nmol/L | Deficiency rate (%) |
| --- | --- | --- | --- |
| **Sex** |  |  |  |
| Male | 4,273 | 40.43 (27.78,55.96) | 66.5 |
| Female | 6,658 | 34.59 (23.39,51.04) | 73.8 |
| *P* value |  | <0.01 | <0.01 |
| **Age group** |  |  |  |
| <30 years | 1,793 | 32.6 (20.7,41.6) | 85.6 |
| 30–40 years | 2,648 | 36.5 (22.7,46.1) | 79.6 |
| 40–50 years | 2,129 | 39.7 (24.7,52.7) | 72.0 |
| 50–60 years | 2,292 | 46.0 (29.4,52.7) | 63.3 |
| 60–70 years | 1,351 | 51.0 (32.5,65.1) | 53.2 |
| >70 years | 718 | 49.8 (31.1,62.5) | 57.7 |
| *P* value (trend) |  | <0.01 | <0.01 |
| **Season** |  |  |  |
| Spring (Mar–May) | 2,765 | 33.9 (20.7,41.5) | 83.5 |
| Summer (Jun–Aug) | 2,529 | 42.1 (24.9,52.7) | 71.9 |
| Autumn (Sep–Nov) | 2,159 | 47.9 (31.3,61.0) | 57.4 |
| Winter (Dec–Feb) | 3,478 | 42.6 (27.2,54.6) | 68.7 |

|  |  |  |
| --- | --- | --- |
| *P* value | <0.01 | <0.01 |

\*Notes: Deficiency defined as 25(OH)D <50 nmol/L. *P* values from Mann-Whitney U test (sex) or Kruskal-Wallis H test (age, season); for deficiency rates, chi-square test.\*

### Age- and sex-specific patterns

In participants aged <50 years, men had significantly higher 25(OH)D levels than women (P < 0.05). No significant sex difference was observed in the ≥50 years age groups (P > 0.05). For example, in the <30 years group, the deficiency rate was 75.6% in men vs. 89.2% in women; in the 30–40 years group, 74.5% vs. 82.4%; in the 50–60 years group, it became comparable (63.5% in men vs. 63.1% in women).

### Seasonal pattern

Autumn showed the highest serum 25(OH)D levels (median 47.9 nmol/L) and the lowest deficiency rate (57.4%). Spring had the lowest levels (median 33.9 nmol/L) and the highest deficiency rate (83.5%). This pattern persisted across almost all age and sex subgroups.

### Machine learning-identified key factors

The LASSO regression path analysis selected four key predictors (age, season, sex, and BMI) as the optimal subset (cross-validated MSE = 0.36 at log(λ) ≈ –5). The optimized Random Forest model achieved a cross-validated RMSE of 21.427.

**SHAP feature importance** (Figure 1) revealed a clear hierarchy:

**Age:** mean SHAP = 5.604

**Season:** mean SHAP = 4.104

**Sex:** mean SHAP = 1.533

**BMI:** mean SHAP = 1.501

**Figure 1.**
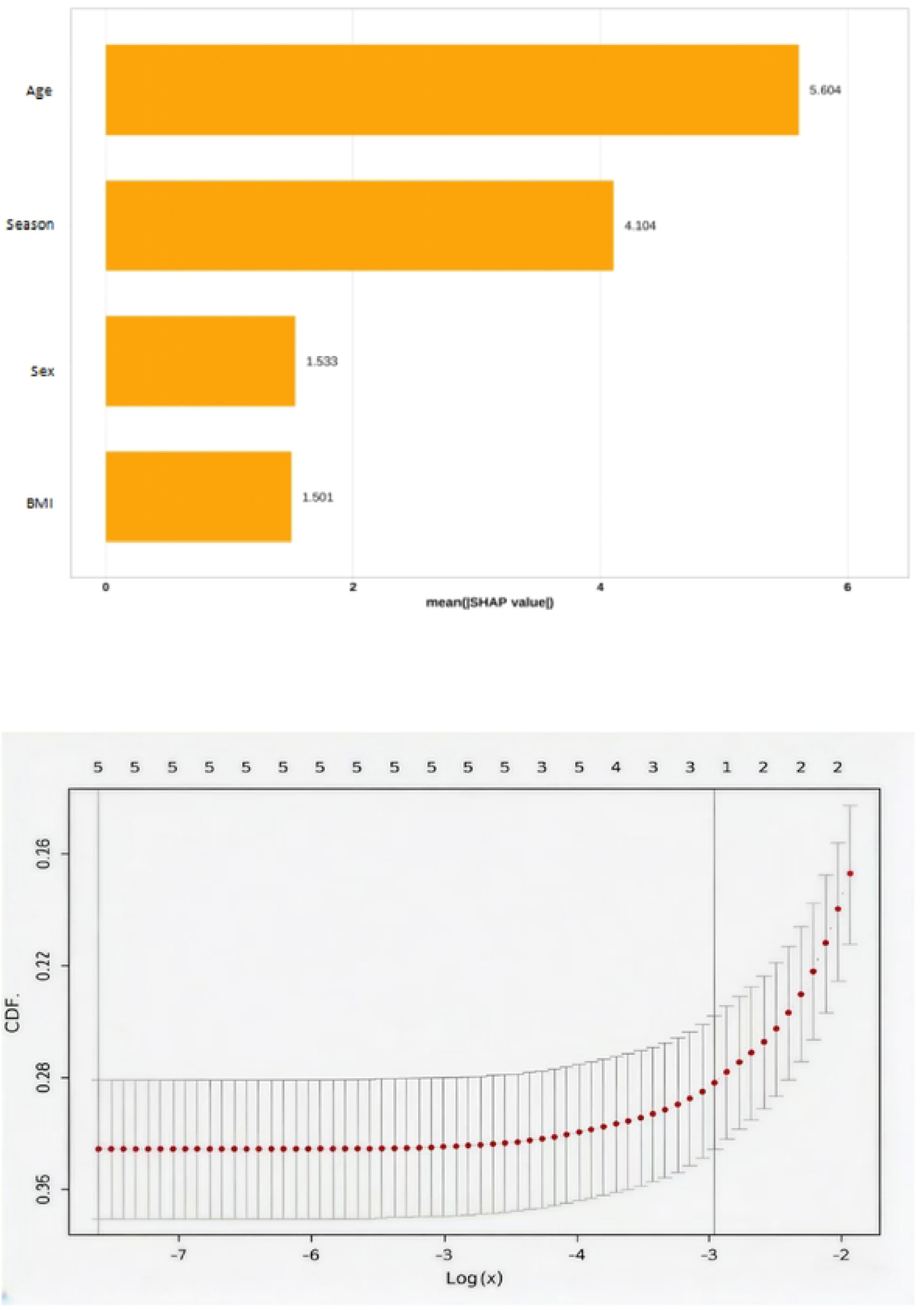
SHAP bar plot of feature importance. The mean absolute SHAP values show that age and season are the dominant predictors of serum 25(OH)D levels.

Age and season together contributed more than 70% of the total importance, far outweighing sex and BMI.

## Discussion

This study reveals an alarmingly high prevalence of vitamin D deficiency (70.98%) among health examination adults in Guiyang. This rate is substantially higher than those reported in Beijing, Kunming, and Lanzhou [8–11]. The phenomenon can be attributed to Guiyang’s unique geographic and climatic setting—low latitude (26°N) combined with high altitude (approximately 1100 m) but persistently cloudy and rainy weather (“no three consecutive sunny days”). Although lower latitude would theoretically increase UV intensity, the frequent cloud cover and limited annual sunshine hours drastically reduce actual UV exposure, hindering sufficient cutaneous vitamin D synthesis [10]. As Holick [6] emphasized, sun exposure remains the major source of vitamin D for most children and adults, and factors such as clothing, sunscreen use, glass, and atmospheric conditions (e.g., cloud cover, ozone absorption) can dramatically reduce or eliminate UVB-driven cutaneous production.Thus, our findings reflect not merely laboratory assays but a pressing public health challenge inherent to such “cloudy highland” environments.

Consistent with most previous studies [9,13,14], men had significantly higher 25(OH)D levels than women. Beyond biological differences, sociocultural and behavioral factors are likely predominant: young women in Guiyang commonly pursue fair skin (“white is beautiful”), work indoors, and diligently use umbrellas, sunscreens, and other photoprotection measures, leading to severely reduced UV exposure [11,15].Holick [6] noted that individuals with increased skin melanin pigmentation and those who practice abstinence from direct sun exposure are at especially high risk for vitamin D deficiency. Furthermore, the proper application of sunscreen with SPF 30 absorbs approximately 97.5% of UVB radiation on the skin surface, thereby reducing cutaneous vitamin D production by a similar magnitude [6]. These mechanisms likely contribute to the particularly high deficiency rate observed in young women in our cohort.

Of particular note, the relationship between vitamin D and age exhibited a “U-shaped” pattern in deficiency risk, with the youngest group (<30 years) being the most severely affected (deficiency rate 85.6%). This group is predominantly composed of office workers with sedentary lifestyles and strong sun-avoidance behaviors.As Holick [6] observed, even in the United States—where milk and some juices are fortified with vitamin D—50% of children aged 1–5 years and 70% of children aged 6–11 years had serum 25(OH)D levels below 30 ng/mL (75 nmol/L), largely due to decreased milk consumption, increased sun protection, and rising obesity rates. The situation is likely worse in regions without mandatory food fortification, such as China. The modest decline in levels among those >70 years likely reflects reduced outdoor activity, limited mobility, and lower cutaneous synthetic capacity [16]. Interestingly, the sex difference disappeared after age 50, which might indicate that postmenopausal women adopt healthier behaviors (increased outdoor activity or active vitamin D supplementation).

The seasonal variation—autumn peak, spring trough—closely matches Guiyang’s climate of “four distinct seasons, damp and rainy spring, crisp autumn.” Autumn offers optimal weather for outdoor activities and sun exposure, whereas spring is characterized by persistent rain, low sunlight, and “late spring cold spells.” This pattern differs from northern Chinese cities where summer peaks have been reported [15,18,19], again highlighting the decisive role of local climate.

It is worth noting that the definition of vitamin D sufficiency remains debated. The Institute of Medicine [2011] considers 20 ng/mL (50 nmol/L) adequate for bone health, whereas the Endocrine Society [6] recommends ≥30 ng/mL (75 nmol/L) for maximum musculoskeletal health. The latter threshold is supported by evidence that serum parathyroid hormone levels begin to plateau when 25(OH)D reaches 30–40 ng/mL, and that the prevalence of secondary hyperparathyroidism and osteomalacia increases substantially below this level [6]. In our study, we adopted the Chinese consensus definition (<50 nmol/L as deficiency), which aligns with the IOM threshold. Even using this relatively conservative cut-off, we observed a 70.98% deficiency rate, indicating that the true prevalence of suboptimal vitamin D status for musculoskeletal health would be even higher if the Endocrine Society’s threshold were applied.

The machine learning framework (Random Forest + SHAP) provided an objective, data-driven confirmation of the traditional findings. The SHAP importance hierarchy (age > season » sex ≈ BMI) robustly demonstrates that age and season are the dominant drivers of vitamin D status in this population.

### Limitations

This study has several limitations. It is a single-center retrospective study, which may limit the generalizability of its findings. We did not exclude individuals with gastrointestinal disorders, thyroid dysfunction, liver or kidney insufficiency, or those taking vitamin D supplements. Detailed baseline characteristics are provided in Supplementary Table S1. Future studies incorporating detailed dietary surveys, direct sunlight exposure measurements, and stricter exclusion of such confounders will provide more precise estimates.

From a public health perspective, Holick (2017) argues that improving vitamin D status can be achieved through sensible sun exposure and increased food fortification, as universal screening is not cost-effective. For populations in cloudy highland regions like Guiyang, where effective UVB exposure is severely limited even during summer months, vitamin D supplementation becomes particularly important. The Endocrine Society recommends 1500–2000 IU daily for adults to prevent and treat deficiency, with obese individuals requiring 2–3 times higher doses. Our findings support the need for region-specific public health strategies, including targeted supplementation for high-risk groups (young adults, females, and the elderly) and seasonal intervention during spring when deficiency is most severe.

## Conclusions

Vitamin D deficiency is highly prevalent and severe among the health examination population in Guiyang. Age and season are the overriding determinants, as objectively quantified by SHAP analysis. Based on these findings, we propose the following public health recommendations:

1. **Targeted screening and health education** – Include serum 25(OH)D testing in routine checkups for young adults, female office workers, and the elderly in this region.
2. **Seasonal intervention strategies** – Emphasize vitamin D monitoring and supplementation during spring, the season with the lowest levels and highest deficiency rate.
3. **Clinical reference** – Local clinicians should maintain a low threshold for assessing vitamin D status, especially in patients with skeletal or metabolic disorders.

## Data Availability

The data that support the findings of this study are derived from health examination records of Guiyang First People's Hospital. This study was approved by the Medical Ethics Committee of Guiyang First People's Hospital (approval date: November 11, 2025). Due to ethical restrictions protecting patient privacy, the raw data are not publicly available. De-identified data may be requested from the corresponding author (Pan Ben-you) upon reasonable scientific request, subject to institutional ethical approval.

## Declaration

Ethics approval and consent to participate Not applicable (retrospective analysis of anonymized data).

Consent for publication Not applicable.

Availability of data and materials The datasets used and/or analyzed during the current study are available from the corresponding author upon reasonable request.

Competing interests The authors declare that they have no competing interests. Funding No funding was received for this study.

Authors’ contributions PBY designed the study, performed the analyses, and drafted the manuscript. WXD contributed to data collection and interpretation. YHL supervised the study and revised the manuscript critically. All authors read and approved the final manuscript.

## Acknowledgment

None.

## Supplementary Materials

**Supplementary Table S1.**
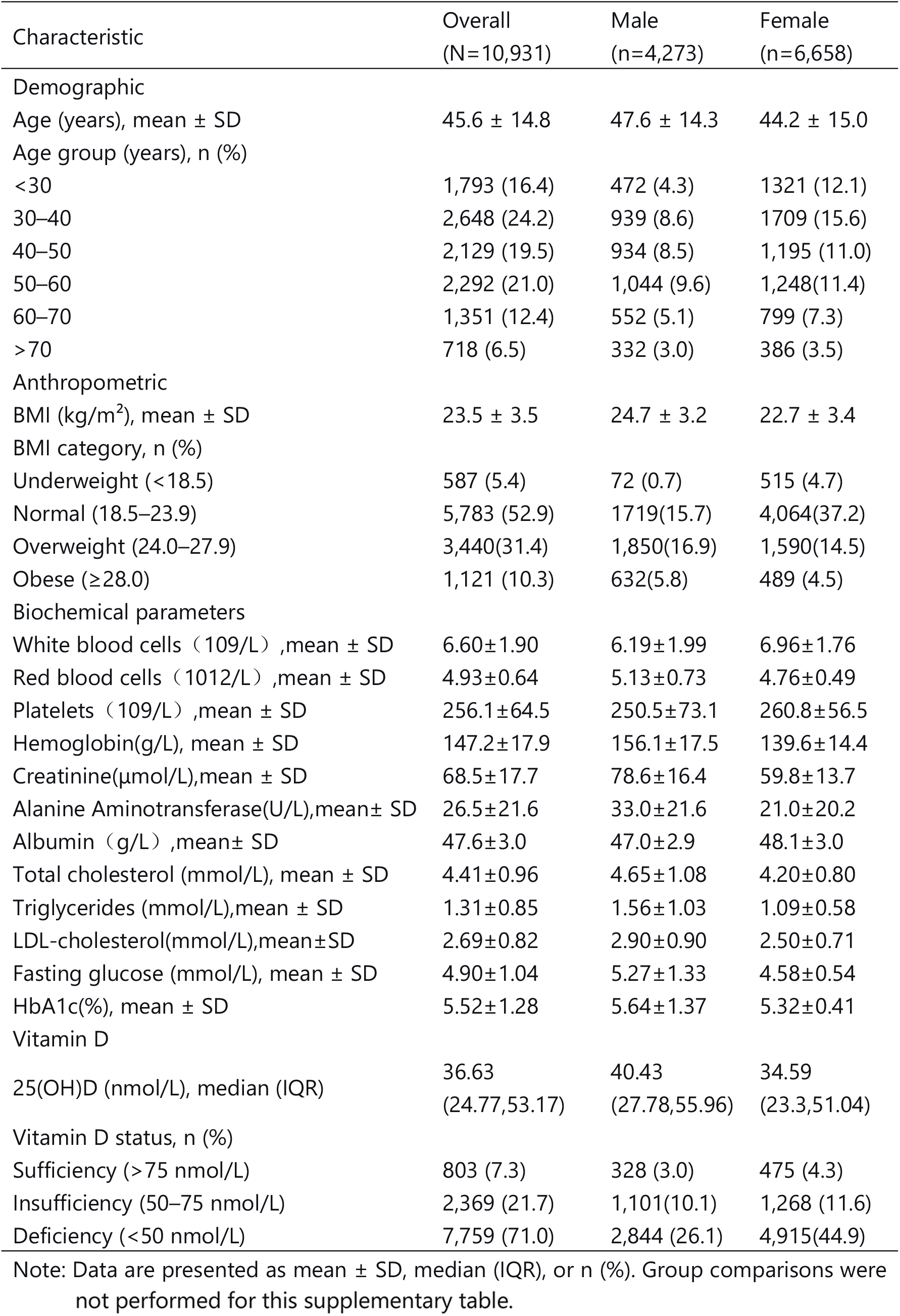
Baseline characteristics of the study population (N = 10,931)

| Characteristic | Overall<br>(N=10,931) | Male<br>(n=4,273) | Female<br>(n=6,658) |
| --- | --- | --- | --- |
| Demographic |  |  |  |
| Age (years), mean ± SD | 45.6 ± 14.8 | 47.6 ± 14.3 | 44.2 ± 15.0 |
| Age group (years), n (%) |  |  |  |
| <30 | 1,793 (16.4) | 472 (4.3) | 1321 (12.1) |
| 30–40 | 2,648 (24.2) | 939 (8.6) | 1709 (15.6) |
| 40–50 | 2,129 (19.5) | 934 (8.5) | 1,195 (11.0) |
| 50–60 | 2,292 (21.0) | 1,044 (9.6) | 1,248 (11.4) |
| 60–70 | 1,351 (12.4) | 552 (5.1) | 799 (7.3) |
| >70 | 718 (6.5) | 332 (3.0) | 386 (3.5) |
| Anthropometric |  |  |  |
| BMI (kg/m<sup>2</sup>), mean ± SD | 23.5 ± 3.5 | 24.7 ± 3.2 | 22.7 ± 3.4 |
| BMI category, n (%) |  |  |  |
| Underweight (<18.5) | 587 (5.4) | 72 (0.7) | 515 (4.7) |
| Normal (18.5–23.9) | 5,783 (52.9) | 1719 (15.7) | 4,064 (37.2) |
| Overweight (24.0–27.9) | 3,440 (31.4) | 1,850 (16.9) | 1,590 (14.5) |
| Obese (≥28.0) | 1,121 (10.3) | 632 (5.8) | 489 (4.5) |
| Biochemical parameters |  |  |  |
| White blood cells ($\times 10^9/L$), mean ± SD | 6.60±1.90 | 6.19±1.99 | 6.96±1.76 |
| Red blood cells ($\times 10^{12}/L$), mean ± SD | 4.93±0.64 | 5.13±0.73 | 4.76±0.49 |
| Platelets ($\times 10^9/L$), mean ± SD | 256.1±64.5 | 250.5±73.1 | 260.8±56.5 |
| Hemoglobin(g/L), mean ± SD | 147.2±17.9 | 156.1±17.5 | 139.6±14.4 |
| Creatinine(μmol/L), mean ± SD | 68.5±17.7 | 78.6±16.4 | 59.8±13.7 |
| Alanine Aminotransferase(U/L), mean± SD | 26.5±21.6 | 33.0±21.6 | 21.0±20.2 |
| Albumin (g/L), mean± SD | 47.6±3.0 | 47.0±2.9 | 48.1±3.0 |
| Total cholesterol (mmol/L), mean ± SD | 4.41±0.96 | 4.65±1.08 | 4.20±0.80 |
| Triglycerides (mmol/L), mean ± SD | 1.31±0.85 | 1.56±1.03 | 1.09±0.58 |
| LDL-cholesterol(mmol/L), mean±SD | 2.69±0.82 | 2.90±0.90 | 2.50±0.71 |
| Fasting glucose (mmol/L), mean ± SD | 4.90±1.04 | 5.27±1.33 | 4.58±0.54 |
| HbA1c(%), mean ± SD | 5.52±1.28 | 5.64±1.37 | 5.32±0.41 |
| Vitamin D |  |  |  |
| 25(OH)D (nmol/L), median (IQR) | 36.63<br>(24.77,53.17) | 40.43<br>(27.78,55.96) | 34.59<br>(23.3,51.04) |
| Vitamin D status, n (%) |  |  |  |
| Sufficiency (>75 nmol/L) | 803 (7.3) | 328 (3.0) | 475 (4.3) |
| Insufficiency (50–75 nmol/L) | 2,369 (21.7) | 1,101 (10.1) | 1,268 (11.6) |
| Deficiency (<50 nmol/L) | 7,759 (71.0) | 2,844 (26.1) | 4,915 (44.9) |

Note: Data are presented as mean ± SD, median (IQR), or n (%). Group comparisons were
not performed for this supplementary table.

